# Sleep problems and sleep quality in the general adult population living in South Tyrol (Italy), a cross-sectional survey study

**DOI:** 10.1101/2024.12.05.24318572

**Authors:** Dietmar Ausserhofer, Giuliano Piccoliori, Adolf Engl, Pasqualina Marino, Verena Barbieri, Stefano Lombardo, Timon Gärtner, Christian J. Wiedermann

**Affiliations:** Institute of General Practice and Public Health, Claudiana, Bolzano-Bozen, Italy; Provincial Institute of Statistics (ASTAT), Bolzano-Bozen, Italy; Claudiana Research, College of Healthcare Professions, Bolzano-Bozen, Italy

**Keywords:** adults, sleep quality, sleep behaviors, cross-sectional study, population-based survey, stratified probabilistic sampling

## Abstract

This cross-sectional, population-based survey study aimed to (1) investigate the prevalence of sleep problems and poor sleep quality and (2) explore the associated sociodemographic and health-related factors. A stratified probabilistic sample of 4,000 adults aged ≥ 18 years living in the Autonomous Province of Bolzano (South Tyrol, Italy) was invited to complete a questionnaire. Sleep quality was assessed using the brief version of the Pittsburgh Sleep Quality Index. Descriptive and logistic regression analyses were performed to analyze the data. A total of 2,090 adults (53%) completed the survey. Poor sleep quality was reported by 17.8%, with 28.2% of participants reporting insufficient sleep duration (i.e., six hours or less), 12.7% having problems staying asleep (i.e. waking up to 3-4 times a week and unable to fall asleep again), and 8.7% having problems getting to sleep (i.e. >30 minutes). Sleep problems and poor sleep quality were associated with sociodemographic and health-related factors, including sex, age, mother tongue, overall health status, chronic disease, and sleep hygiene. Italian-speaking participants reported poorer sleep quality and greater difficulty staying asleep compared to German-speaking participants, highlighting potential sociocultural influences on sleep health. Sleep quality and other problems are prevalent among adults in South Tyrol. Further research on public health interventions that promote sleep health literacy and hygiene is needed.

## Introduction

Sleep is a fundamental biological process essential for human health and well-being [1, 2]. Sleep health–that is, adequate sleep quality and quantity–is crucial for maintaining physical and mental health, cognitive function, and overall quality of life [3, 4]. To date, there is a vast amount of evidence that sleep disorders and poor sleep quality are associated with physical health outcomes, including poor cardiovascular health, high blood pressure, obesity, and diabetes [5–8]; mental health consequences such as depression, anxiety, and suicidal behavior [9–11]; and overall mortality [12, 13]. Moreover, the negative impact of sleep problems extends beyond individual health, affecting workplace productivity [14], healthcare costs [15], and public safety [16].

According to the American Academy of Sleep Medicine and Sleep Research Society 7– 8h of total sleep is recommended as an adequate sleep quantity for the adult population [17]. Sleep quality is defined as an individual’s self-satisfaction with all aspects of their sleep experience [18]. The US National Sleep Foundation has put forward the four key indicators for good sleep quality: (i) less difficulty in initiating sleep, (ii) sleeping more time while in bed, (iii) less than two instances of night waking and (iv) ability to go back to sleep within 20 min of a night awakening [19].

Sleep problems and poor sleep quality have become increasingly prevalent in modern society and affect a significant proportion of the global population [1]. Between 10% and 30% of the general adult population are affected by sleep problems and poor sleep quality [20, 21], with a prevalence of diagnosed sleep disorders reported in the literature ranging between 5% and 15% [22–24]. The COVID-19 pandemic has had a significant impact on sleep quality and disturbances worldwide. A meta-analysis of 63 studies revealed a 40% higher risk of poor sleep quality during the pandemic than pre-pandemic [25], with increased stress and anxiety related to health concerns, economic uncertainty, social isolation, working from home, reduced physical activity, increased screen time, and COVID-19 infection itself as influential factors. Sociodemographic (e.g., women [26, 27], older adults [28], economic status, race/ethnicity [29, 30]), health-related (chronic diseases and multimorbidity [31, 32]), and environmental factors [30] are associated with sleep disorders and poor sleep quality.

While sleep research has been conducted in various populations worldwide [1], there is still limited information on sleep problems and sleep quality in Italy [33–35]. The most recent cross-sectional study, conducted in 2019, including a sample of 3120 subjects from the Italian general population, found that 14.2% of Italian adults were dissatisfied with sleep and 29.5% reported insufficient sleep duration [35]. South Tyrol, an autonomous province in northern Italy, has a unique cultural and linguistic landscape, with influences from both Italian and German-speaking populations. However, no data on sleep problems or sleep quality are currently available.

Given the importance of sleep in health, it is vital to monitor the prevalence of sleep quality and sleep problems at the population level and compare these estimates and trends within and between countries [1]. However, it remains unclear how sleep quality and sleep problems have developed in the general population of Italy and other countries after the COVID-19 pandemic. South Tyrol, an autonomous province in northern Italy, is characterised by a unique linguistic and cultural landscape, with approximately 70% of the population speaking German and 25% speaking Italian. The coexistence of two linguistic groups within the same geographical region presents a valuable opportunity to investigate health-related behaviours, including sleep quality, in a population influenced by diverse cultural factors.

Examining sleep patterns in this context can contribute to a more comprehensive understanding of sleep health in diverse populations, and inform public health initiatives to improve sleep health tailored to multilingual and multicultural populations.

Therefore, this study aimed to (1) describe the prevalence of poor sleep quality and sleep problems among adults living in the culturally distinct Autonomous Province of Bolzano (South Tyrol, Italy), and (2) explore the sociodemographic and health-related factors associated with sleep quality, with a specific focus on the potential influence of linguistic and cultural differences between German- and Italian-speaking groups.

## Methods

### Study Design

This cross-sectional, population-based survey was conducted jointly by the Provincial Institute of Statistics (ASTAT; Istituto Provinciale di Statistica – Landesinstitut für Statistik) and the Institute of General Practice and Public Health in the Autonomous Province of Bolzano, South Tyrol, between 1 March and 30 May 2024.

### Setting and sample

South Tyrol, the Autonomous Province of Bolzano, is part of the Trentino–Alto Adige region in Italy, next to Austria (total population: 534.912), with approximately 70% German-speaking, 25% Italian-speaking, and 5% other languages. The target population of the survey was approximately 400,000 individuals aged 18 years and above residing in South Tyrol.

Stratified probabilistic sampling is used in this study. The ASTAT randomly selected adults aged ≥ 18 years, stratified by age (18-34, 35-54, 55, and above), sex (male and female), citizenship (Italian or other), and residence (municipalities), from the register of the current resident population in the whole province. To ensure an adequate level of precision, 4,000 individuals were sampled considering the distribution of and variation between the strata.

### Participant survey

The participant survey was designed collaboratively by the ASTAT and the Institute of General Practice and Public Health. The German and Italian versions, translated from ASTAT, were reviewed for language equivalence by a research group at the Institute for General Practice and Public Health.

*Sleep quality* was measured using the brief version of the Pittsburgh Sleep Quality Index (B-PSQI) [36]. Although polysomnography, actigraphy, or movements captured using other phone apps or body wearables are objective measurements of sleep quality, they are too expensive and impede data collection in large samples. Thus, most epidemiological studies on sleep quality rely on self-reported measures of sleep, such as the Pittsburgh Sleep Quality Index (PSQI) [37]. The original version of the PSQI is a widely used self-report questionnaire designed to measure sleep quality and disturbances over a one-month period [37]. It has been extensively employed in both clinical and research settings and demonstrated reliability and validity across diverse populations and conditions [38, 39]. The original version consists of 19 items grouped into seven component scores. We used the brief version described and tested by Sancho-Domingo et al. (2021) [36], who provided evidence based on internal validity by applying confirmatory factor analyses and sensitivity and specificity for classifying poor sleepers, such as the full PSQI version. The brief version uses six items from the original version to assess five dimensions: perceived sleep quality, sleep duration, sleep efficiency, sleep latency, and sleep disturbances. Thus, the six questions of the B-PSQI yielded five components rated on a scale from 0 to 3, similar to the original version, with higher scores indicating greater sleep disturbance or poorer sleep quality. The component scores were summed to produce a global B-PSQI score ranging from 0 to 15, with scores above 5 indicating poor sleep quality [36]. In our study, we used six items from the original Italian [40] and German version [20] of the PSQI. Reliability testing revealed acceptable internal consistency for both language versions of the B-PSQI(i.e., Cronbach’s α = 0.74 for the German version; Cronbach’s α = 0.77 for the Italian version).

*Participants’ sociodemographic characteristics* included age (birth year), gender (male/female), native tongue (German/Italian/Ladin or Others), citizenship (Italy/other country), educational level (below school/high school or higher), community and region of origin (rural/urban), and living situation (alone/with spouse or family member or with parents or children). *Health-related factors* included self-reported health status (poor, moderate/good, or very good), diagnosed chronic disease (none or nine types of chronic disease), adequate sleep (never, seldom/often, or always), and intake of sleep medications (never/once a week or more often).

### Data collection

Letters were mailed from the ASTAT to randomly sampled participants to inform them about the study and invite them to voluntarily participate by completing the survey alone or with the aid of a family member. The survey was completed through online self-completion or telephone interviews with collaborators from the ASTAT. One month after the first letter, a second letter was sent to inform participants about the study and invite them to participate. An online survey was created using LimeSurvey [41].

### Ethical aspects

Ethical approval was obtained from the Institutional Board of the Institute of General Practice and Public Health, Bolzano, Italy. All study procedures were in accordance with the 1964 Helsinki Declaration and its amendments, European Union General Data Protection Regulation (679/2016), and Italian Data Protection Law (196/2003). Before completing the online questionnaire, participants were asked to provide informed consent. Filling out the paper questionnaire and sending it back by post were considered participants’ informed consent. Participation in this study was voluntary. All data from study participants were anonymized to protect their identities.

### Statistical analysis

Only fully completed questionnaires were included in the statistical analyses. Descriptive statistics (e.g. frequency) were calculated to describe the measured variables. To adjust for nonresponse bias and ensure that the sample was representative of the target population, weighted descriptive statistics were calculated using post-stratification weights to replicate the distribution of the population according to age, gender, citizenship, and residence. For continuous variables, weighted medians and interquartile ranges were computed; for categorical variables, weighted proportions and 95% confidence intervals were reported. To explore the association between sociodemographic and health-related factors and poor sleep quality, insufficient sleep duration, sleep latency problems getting asleep, and problems staying asleep, four multivariate binary logistic regression models applying generalized linear modelling were computed. Multicollinearity testing using the variance inflation factor (VIF) revealed no evidence of collinearity among the independent variables (VIF for all variables < 1.4). All analyses were performed using the R Statistical Software (v4.4.2) [42] in the RStudio environment (2024.09.1+394) [43] and the packages tidyr [44], survey [45] and lme4 [46]. A *P* value of less than 0.05 was considered significant.

## Results

### Sample characteristics

In total, 2,090 adults from the general population completed the survey, reflecting a response rate of 53%. As described in Table 1, the majority of participants were female (55.2%), 55 years of age or above (50.7%), German-speaking (66.9%), living with partners/families (81.7%), and had the highest educational level below high school (55.7%). Overall, 61.9% reported good or very good health and 61.3% had no diagnosed chronic disease. Among the participating adults, 80.8% often or always cared for enough sleep and 82.9% never took sleep medications (as natural supplements or by prescription) over the last month (see Table 1).

**Table 1.** Characteristics of the adults participating in the study (N=2,090)

| Characteristics | n | % |
| --- | --- | --- |
| **Gender - % (n)** |  |  |
| Female | 1153 | 55.2 |
| Male | 937 | 44.8 |
| **Age (years) - % (n)** |  |  |
| 18-34 | 383 | 18.3 |
| 35-54 | 647 | 31.0 |
| 55-99 | 1060 | 50.7 |
| **Native tongue - % (n)** |  |  |
| German | 1398 | 66.9 |
| Italian | 499 | 23.9 |
| Ladin or Other | 193 | 9.2 |
| **Citizenship - % (n)** |  |  |
| Italian | 2011 | 96.2 |
| Other | 79 | 3.8 |
| **Community - % (n)** |  |  |
| Urban | 381 | 18.2 |
| Rural | 1709 | 81.8 |
| **Living situation - % (n)** |  |  |
| Alone | 383 | 18.3 |
| With partner/family | 1326 | 63.4 |
| With children | 785 | 37.6 |
| With parents | 196 | 9.4 |
| With other family members | 124 | 5.9 |
| **Educational level - % (n)** |  |  |
| Middle school or lower | 492 | 23.5 |
| Vocational school (2-3 years) | 674 | 32.2 |
| High School (5 years) | 530 | 25.4 |
| University | 394 | 18.9 |
| **Health status – 0-100** |  |  |
| Poor or moderate | 796 | 38.1 |
| Good or very good | 1294 | 61.9 |
| **Chronic disease** |  |  |
| None | 1282 | 61.3 |
| Lung diseases (e.g., asthma, chronic obstructive pulmonary disease) | 97 | 4.6 |
| Cardiovascular diseases (e.g., coronary artery disease, atrial fibrillation, heart failure, stroke/cerebral circulatory disorders) | 171 | 8.2 |
| High blood pressure | 399 | 19.1 |
| Kidney diseases | 43 | 2.1 |
| Immune system disorders | 73 | 3.5 |
| Cancer | 93 | 4.4 |
| Metabolic disorders (e.g., diabetes, overweight/obesity) | 157 | 7.5 |
| Liver diseases | 24 | 1.1 |
| Mental disorders (e.g., depression, anxiety) | 122 | 5.8 |
| **Taking care for enough sleep** |  |  |
| Never or seldom | 401 | 19.2 |
| Often or always | 1689 | 80.8 |
| **Intake of sleep medications during last month (natural supplements or by prescription)** |  |  |
| Never | 1732 | 82.9 |
| Once a week or more often | 358 | 17.1 |

### Sleep quality and sleep problems

Of the 2,090 participants, 17.9% (95%CI: 16.2-19.4) described their overall sleep quality as quite bad or very bad. While the median sleep duration was 7.0 h per night (IQR: 2.0; data not shown in tables), insufficient sleep duration, i.e., six hours or less, was reported from 28.2% (95%CI: 26.3-30,1). A total of 12.7% (95%CI: 11.3-14.1) reported having problems staying asleep, that is, waking up 3-4 times a week and cannot fall asleep again, and 8.7% (95%CI: 7.5-9.9) reported having problems getting to sleep, that is, >30 minutes.

### Socio-demographic and health-related factors associated with poor sleep quality and sleep problems

Multivariate logistic regression analysis revealed several factors associated with poor sleep quality and sleep problems, such as insufficient sleep duration, problems getting to sleep, problems staying asleep, and other problems (Tables 2 and 3).

**Table 2.** Multivariate logistic regression: factors associated with poor sleep quality and insufficient sleep duration (6 hours or less) of adults living in South Tyrol (N=2,090)

|  | Poor sleep quality |  |  | Insufficient sleep duration |  |  |
| --- | --- | --- | --- | --- | --- | --- |
|  | OR° | 95%CI | P | OR | 95%CI | P |
| Females (reference group: males) | 2.03 | 1.63, 2.55 | <0.001 | 1.96 | 1.48, 2.61 | <0.001 |
| Age (reference group: 18-34 years) |  |  |  |  |  |  |
| 35-54 years | 1.10 | 0.78, 1.56 | 0.6 | 1.31 | 0.82, 2.15 | 0.3 |
| 55-99 years | 1.69 | 1.20, 2.39 | 0.003 | 1.72 | 1.09, 2.79 | 0.023 |
| Native tongue (reference group: German) |  |  |  |  |  |  |
| Italian | 1.40 | 1.05, 1.86 | 0.023 | 1.30 | 0.91, 1.84 | 0.15 |
| Ladin & others | 1.12 | 0.75, 1.66 | 0.6 | 1.05 | 0.61, 1.72 | 0.9 |
| Rural community (reference group: urban community) | 1.11 | 0.81, 1.52 | 0.5 | 1.09 | 0.75, 1.62 | 0.7 |
| Living alone (reference group: living with family) | 1.04 | 0.79, 1.37 | 0.8 | 0.76 | 0.53, 1.08 | 0.14 |
| Educational level high school or higher (reference group: below high school) | 0.77 | 0.60, 0.97 | 0.027 | 0.60 | 0.44, 0.81 | <0.001 |
| Good or very good health status (reference: poor or moderate) | 0.51 | 0.41, 0.65 | <0.001 | 0.54 | 0.41, 0.72 | <0.001 |
| Chronic disease (reference group: none) | 1.31 | 1.03, 1.67 | 0.027 | 1.58 | 1.18, 2.14 | 0.003 |
| Often or always taking care for enough sleep (reference group: never or seldom) | 0.18 | 0.14, 0.23 | <0.001 | 0.26 | 0.20, 0.35 | <0.001 |
| Use of sleep medication (reference group: none) | 2.27 | 1.74, 2.95 | <0.001 | 1.67 | 1.22, 2.27 | 0.001 |

OR: Odds Ratio, CI: Confidence Interval

**Table 3.** Multivariate logistic regression: factors associated with having problems staying asleep (i.e., waking up 3-4 times a week and can’t falling asleep again) and problems getting to sleep of adults living in South Tyrol (N=2,090)

|  | Problems getting to sleep |  |  | Problems staying asleep |  |  |
| --- | --- | --- | --- | --- | --- | --- |
|  | OR | 95%CI | P | OR | 95%CI | P |
| Females (reference group: males) | 1.91 | 1.37, 2.69 | <0.001 | 1.07 | 0.86, 1.33 | 0.6 |
| Age (reference group: 18-34 years) |  |  |  |  |  |  |
| 35-54 years | 0.42 | 0.26, 0.67 | <0.001 | 1.50 | 1.07, 2.13 | 0.021 |
| 55-99 years | 0.60 | 0.38, 0.94 | 0.024 | 1.90 | 1.35, 2.70 | <0.001 |
| Native tongue (reference group: German) |  |  |  |  |  |  |
| Italian | 0.79 | 0.50, 1.23 | 0.3 | 1.62 | 1.22, 2.16 | <0.001 |
| Ladin & others | 1.82 | 1.10, 2.95 | 0.017 | 1.04 | 0.69, 1.55 | 0.8 |
| Rural community (reference group: urban community) | 1.50 | 0.93, 2.52 | 0.11 | 0.95 | 0.70, 1.29 | 0.7 |
| Living alone (reference group: living with family) | 1.05 | 0.69, 1.55 | 0.8 | 1.34 | 1.02, 1.76 | 0.034 |
| Educational level (reference group: below high school) | 0.77 | 0.54, 1.09 | 0.15 | 1.08 | 0.85, 1.37 | 0.5 |
| Good or very good health status (reference: poor or moderate) | 0.50 | 0.35, 0.70 | <0.001 | 0.76 | 0.60, 0.96 | 0.024 |
| Chronic disease (reference group: none) | 1.15 | 0.80, 1.65 | 0.4 | 1.14 | 0.89, 1.45 | 0.3 |
| Often or always taking care for enough sleep (reference group: never or seldom) | 0.33 | 0.24, 0.47 | <0.001 | 0.09 | 0.07, 0.12 | <0.001 |
| Use of sleep medication (reference group: none) | 2.03 | 1.41, 2.90 | <0.001 | 1.37 | 1.03, 1.81 | 0.028 |

OR: Odds Ratio, CI: Confidence Interval

Compared to males, females had nearly two times higher probability of reporting poor sleep quality (OR: 2.03, 95% CI: 1.63–2.55, *P* < 0.001), insufficient sleep duration (OR: 1.96, 95% CI: 1.48–2.61, *P* < 0.001), and problems getting to sleep (OR: 1.91, 95% CI: 1.37–2.69, *P*< 0.001), but not problems staying asleep. Older adults (55 years and above) were more likely to report poor sleep quality (OR: 1.69, 95% CI: 1.20–2.39, *P* = 0.003), insufficient sleep duration (OR: 1.72, 95% CI: 1.09–2.79, *P* = 0.023), and problems staying asleep (OR: 1.90, 95% CI: 1.35–2.70, *p* < 0.001) compared to the youngest age group (18–34 years). Both adults aged 35–54 years (OR: 0.42, 95%CI: 0.26-0.57, *P* < 0.001) and adults aged 55 years or older (OR: 0.60, 95%CI: 0.38-0.94, *P* = 0.024) had lower odds of getting to sleep than those aged between 18 and 34 years. Italian-speaking adults were more likely to report poor sleep quality (OR: 1.40, 95% CI: 1.05–1.86, *P* = 0.023) and problems staying asleep (OR, 1.62; 95% CI: 1.22–2.16, *P* < 0.001). compared to German speakers, but this was not significant for insufficient sleep duration and sleep problems. Higher education (high school or higher) reduced the odds of poor sleep quality (OR: 0.77, 95% CI: 0.60–0.97, *P* = 0.027) and insufficient sleep duration (OR: 0.60, 95% CI: 0.44–0.81, *P* < 0.001), yet not for problems getting to sleep and staying asleep.

Adults reporting good or very good health were less likely to report poor sleep quality (OR: 0.51, 95% CI: 0.41–0.65, *P* < 0.001), insufficient sleep duration (OR: 0.54, 95% CI: 0.41–0.72, *P* < 0.001), problems getting to sleep (OR: 0.50, 95% CI: 0.35–0.70, *P* < 0.001), and staying asleep (OR: 0.76, 95% CI: 0.60–0.96, *P* = 0.024) compared to adults with poor or moderate health status. Adults with a diagnosed chronic disease had higher odds of poor sleep quality (OR: 1.31, 95% CI: 1.03–1.67, *P* = 0.027) and insufficient sleep duration (OR: 1.58, 95% CI: 1.18–2.14, *P* = 0.003), yet not problems getting to sleep and staying asleep. Often or always taking care for enough sleep was significantly associated with lower odds for poor sleep quality (OR: 0.18, 95% CI: 0.14–0.23, *P* < 0.001), insufficient sleep duration (OR: 0.26, 95% CI: 0.20–0.35, *P* < 0.001), problems getting to sleep (OR: 0.33, 95% CI: 0.24–0.47, *P* < 0.001) and staying asleep (OR: 0.09, 95% CI: 0.07–0.12, *P* < 0.001). Sleep medication use once a week or more often over the last month was significantly associated with higher odds of poor sleep quality (OR: 2.27, 95% CI: 1.74–2.95, *P* < 0.001), insufficient sleep duration (OR: 1.67, 95% CI: 1.22–2.27, *P* = 0.001), problems getting to sleep (OR: 2.03, 95% CI: 1.41–2.90, *P* < 0.001), and staying asleep (OR: 1.37, 95% CI: 1.03–1.81, *P* = 0.028).

## Discussion

In this cross-sectional study, we aimed to describe the prevalence of poor sleep quality and sleep problems among adults aged ≥ 18 years in South Tyrol, Italy and to explore the associated sociodemographic and health-related factors. The findings highlight that 17.9% of the participants reported poor sleep quality, with insufficient sleep duration being the most prevalent sleep problem (28.2%). Sleep-onset and sleep-maintenance problems affected a notable proportion of participants, with 12.7% reporting difficulty staying asleep and 8.7% experiencing trouble falling asleep. Sleep problems and poor sleep quality were associated with several sociodemographic characteristics, such as sex, age, native tongue, educational level, and health-related factors, including health status, chronic disease, taking care for sufficient sleep, and intake of sleep medications.

One in six participants reported poor sleep quality and over a quarter experienced insufficient sleep duration. Our results align with previous findings from Italy [35], as well as global estimates of sleep problems and poor sleep quality in the general adult population pre-pandemic [20, 21]. In the absence of data on sleep problems and sleep quality post-pandemic, our findings suggest that after an increase in the prevalence of sleep disorders during COVID-19, sleep health might have “normalized” to pre-pandemic levels. However, the substantial prevalence of poor sleep quality in South Tyrol warrants targeted interventions. There is a need to promote sleep health in public health agendas across the globe [1], including sleep health educational programs and awareness campaigns; increasing, standardizing, and centralizing data on sleep quantity and quality; and developing and implementing sleep health policies across sectors of society. Future research is needed to evaluate the outcomes and influences of such campaigns in improving sleep hygiene and literacy, as well as in the diagnosis and treatment of sleep disorders, and to reduce the onset and severity of comorbidities associated with disordered sleep [47].

Females disproportionately reported poor sleep quality, insufficient sleep duration, and problems with sleep initiation, consistent with previous research highlighting gender differences in sleep patterns [26, 27]. While hormonal fluctuations, caregiving roles, and higher stress levels may explain these disparities, more research, from basic science to clinical research, is needed to gain a better understanding of the differences in sleep disorders between men and women in terms of prevention, clinical signs, treatment approaches, prognosis, and psychological and social impacts, to shape future therapeutic strategies [48]. Older adults (≥55 years) were more likely to report poor sleep quality, insufficient sleep, and problems staying asleep. As aging is associated with natural changes in sleep architecture [49], sleep problems and poor sleep quality are often under-recognized and undertreated in older adults, leading to negative outcomes, including falls, depression and anxiety, cognitive impairment, institutionalization, and mortality [50]. Intriguingly, younger adults (18–34 years) had the highest likelihood of experiencing difficulty initiating sleep, potentially due to lifestyle factors such as screen time and irregular sleep schedules [51]. Language and cultural background were also significant factors. Italian speakers were more likely to report poor sleep quality and difficulty staying asleep than German speakers. As little is known about disparities in sleep problems and quality between different ethnicities [52], our findings might reflect some social and cultural variations between these two groups (e.g. later dinner time and thus bedtime among Italian speakers). However, further research is needed to explore these disparities and their implications in sleep health interventions in this region.

Good or very good self-reported health status was consistently associated with better sleep outcomes, whereas chronic diseases increased the likelihood of poor and insufficient sleep. This finding underscores the bidirectional relationship between sleep and health, where poor health may impair sleep, and vice versa [53, 54]. Sleep medication use was linked to increased odds of all measured sleep problems, suggesting that, while medications may address acute issues, they are not a sustainable solution for chronic sleep disturbances[55]. Individuals using sleep medications reported poor sleep quality and ongoing sleep issues, consistent with research indicating that such medications provide short-term relief but fail to address the root causes of chronic sleep disturbances. Tolerance, poor sleep hygiene, or a lack of behavioral interventions may contribute to these outcomes. This underscores the need for a comprehensive approach that integrates pharmacological treatments with non-pharmacological strategies such as cognitive-behavioral therapy for insomnia. Public health campaigns should raise awareness about the limitations of sleep medications and advocate evidence-based alternatives to enhance long-term sleep health.

A recent investigation on the use of sedative psychotropic medications from 2019 to 2023 revealed lower benzodiazepine use in South Tyrol than the national Italian average, with a trend toward increased sedative antidepressant use, especially mirtazapine, which likely reflects regional prescription preferences. Z-drug use was similar across both regions, whereas melatonin exhibited a gradual, albeit lower, increase in the South Tyrol [56].

Encouragingly, individuals who often or always prioritized getting enough sleep reported significantly fewer sleep problems. This finding emphasizes the importance of sleep hygiene practices and reinforces the potential of behavioral and lifestyle interventions, such as education on sleep health literacy and hygiene, to mitigate sleep disturbances [57].

### Implications for Public Health

Our findings underscore the need for and offer valuable insights into potential public health strategies to improve sleep in the South Tyrolean population. Interventions tailored to different cultural groups should address the specific needs of high-risk groups, such as women, older adults, and individuals with chronic illnesses. Public health campaigns that promote sleep health literacy and hygiene may be beneficial [58]. Collaborative campaigns, such as the “Sleep Well, Be Well” from the American Academy of Sleep Medicine, Centers for Disease Control and Prevention and the Sleep Research Society provide interesting examples (https://sleepeducation.org/healthy-sleep/). Within healthcare, general practitioners are well positioned to recognize persistent sleep problems, diagnose sleep disorders, initiate treatments, or refer to sleep specialists [55]. Despite evidence that non-pharmacological treatment is superior in the long-term management of sleep disorders, hypnotics are often considered by general practitioners as the most successful treatment, especially in frail older adults [59]. Therefore, additional efforts to reduce reliance on sleep medications and explore non-pharmacological treatments such as digital cognitive-behavioural therapy for insomnia (dCBT-I) could improve long-term sleep health in the general adult population.

### Strenghts and Limitations

This study benefited from a large representative sample and the inclusion of diverse demographic and health-related variables. However, this study has some limitations that need to be considered. First, its cross-sectional design limits our ability to establish causality or track changes in sleep quality and problems over time. Future longitudinal studies and objective assessments such as actigraphy should provide deeper insights into sleep patterns and their determinants. This study was conducted in South Tyrol, Italy, which may limit the generalizability of our findings to other regions or countries with different health care systems. Another limitation of our study was that we did not assess the potential consequences of poor sleep quality such as sleepiness and alertness, quality of life, impairment at work or school, or impaired interpersonal function.

## Conclusion

Sleep quality and problems are prevalent among adults in South Tyrol, with significant variations based on sex, age, health status, and sleep behaviors. Public health strategies that promote sleep hygiene and address the needs of vulnerable populations are essential to improve sleep health literacy in the region. Further longitudinal research is warranted to monitor the prevalence of sleep problems and sleep quality at the population level over time and to assess the outcomes and influences of public health campaigns in improving sleep quality and health, as well as the diagnosis and treatment of somnipathies and in reducing the onset and severity of comorbidities associated with disordered sleep.

## Data Availability

Data cannot be shared publicly because of legal restrictions, yet are available from the Provincial Institute of Statistics (ASTAT) upon formal request.

## Declaration

### Availability of data and material

Data cannot be shared publicly because of legal restrictions. Anonymized data are available from the Provincial Institute of Statistics (ASTAT) upon request.

## Acknowledgment

NA

## Authors’ contributions

DA, GP, AE, SL, TG, and CJW developed the idea for this study. DA, GP, AE, SL, TG, VB, PM, and CJW contributed to concept, design, and data collection. DA, GP, VB, and CJW contributed to data analysis and interpretation. DA, and CJW drafted the manuscript. All authors contributed to critical revision of the manuscript and approved the final version.

## Literature

1. Lim DC, Najafi A, Afifi L, Bassetti CLA, Buysse DJ, Han F, et al. The need to promote sleep health in public health agendas across the globe. The Lancet Public Health. 2023;8(10):e820–e6. doi: 10.1016/S2468-2667(23)00182-2.

2. Lloyd-Jones DM, Allen NB, Anderson CAM, Black T, Brewer LC, Foraker RE, et al. Life’s Essential 8: Updating and Enhancing the American Heart Association’s Construct of Cardiovascular Health: A Presidential Advisory From the American Heart Association. Circulation. 2022;146(5):e18–e43. doi: doi:10.1161/CIR.0000000000001078.

3. Raha W, Ryan Tak Chun W, Ji-Eun P, Si Woo L, Dinayinie Ekanayake M, Zhigang L, et al. Sleep duration, chronotype, health and lifestyle factors affect cognition: a UK Biobank cross-sectional study. BMJ Public Health. 2024;2(1):e001000. doi: 10.1136/bmjph-2024-001000.

4. Palmer CA, Bower JL, Cho KW, Clementi MA, Lau S, Oosterhoff B, et al. Sleep loss and emotion: A systematic review and meta-analysis of over 50 years of experimental research. Psychological Bulletin. 2024;150(4):440–63. doi: 10.1037/bul0000410.

5. Cespedes Feliciano EM, Quante M, Rifas-Shiman SL, Redline S, Oken E, Taveras EM. Objective Sleep Characteristics and Cardiometabolic Health in Young Adolescents. Pediatrics. 2018;142(1). doi: 10.1542/peds.2017-4085.

6. Saz-Lara A, Lucerón-Lucas-Torres M, Mesas AE, Notario-Pacheco B, López-Gil JF, Cavero-Redondo I. Association between sleep duration and sleep quality with arterial stiffness: A systematic review and meta-analysis. Sleep Health. 2022;8(6):663–70. doi: 10.1016/j.sleh.2022.07.001.

7. Lo K, Woo B, Wong M, Tam W. Subjective sleep quality, blood pressure, and hypertension: a meta-analysis. J Clin Hypertens (Greenwich). 2018;20(3):592–605. Epub 20180219. doi: 10.1111/jch.13220. PubMed PMID: 29457339; PubMed Central PMCID: PMCPMC8031314.

8. Fatima Y, Doi SAR, Mamun AA. Sleep quality and obesity in young subjects: a meta-analysis. Obesity Reviews. 2016;17(11):1154–66. doi: 10.1111/obr.12444.

9. Blanchard AW, Rufino KA, Nadorff MR, Patriquin MA. Nighttime sleep quality & daytime sleepiness across inpatient psychiatric treatment is associated with clinical outcomes. Sleep Medicine. 2023;110:235–42. doi: 10.1016/j.sleep.2023.08.011.

10. Slanitz C, Fuchshuber J, Fink A, Unterrainer H-F. Anxious and depressive symptoms mediate the influence of sleep quality on suicidality in young adults. Frontiers in Public Health. 2024;12. doi: 10.3389/fpubh.2024.1322069.

11. Pereira T, Martins S, Fernandes L. Sleep duration and suicidal behavior: A systematic review. European Psychiatry. 2017;41(S1):s854-s. Epub 2020/03/23. doi: 10.1016/j.eurpsy.2017.01.1699.

12. Cribb L, Sha R, Yiallourou S, Grima NA, Cavuoto M, Baril AA, et al. Sleep regularity and mortality: a prospective analysis in the UK Biobank. Elife. 2023;12. Epub 20231123. doi: 10.7554/eLife.88359. PubMed PMID: 37995126; PubMed Central PMCID: PMCPMC10666928.

13. Yang L, Xi B, Zhao M, Magnussen CG. Association of sleep duration with all-cause and disease-specific mortality in US adults. Journal of Epidemiology and Community Health. 2021;75(6):556–61. doi: 10.1136/jech-2020-215314.

14. Åkerstedt T, Eriksson J, Freyland S, Widman L, Magnusson Hanson LL, Miley- Åkerstedt A. Changes in Sleep Quality, Sleep Duration, and Sickness Absence: A Longitudinal Study with Repeated Measures. Healthcare (Basel). 2024;12(14). Epub 20240711. doi: 10.3390/healthcare12141393. PubMed PMID: 39057537; PubMed Central PMCID: PMCPMC11275330.

15. Huyett P, Bhattacharyya N. Incremental health care utilization and expenditures for sleep disorders in the United States. J Clin Sleep Med. 2021;17(10):1981–6. doi: 10.5664/jcsm.9392. PubMed PMID: 33949943; PubMed Central PMCID: PMCPMC8494101.

16. Gottlieb DJ, Ellenbogen JM, Bianchi MT, Czeisler CA. Sleep deficiency and motor vehicle crash risk in the general population: a prospective cohort study. BMC Med. 2018;16(1):44. Epub 20180320. doi: 10.1186/s12916-018-1025-7. PubMed PMID: 29554902; PubMed Central PMCID: PMCPMC5859531.

17. Watson NF, Badr MS, Belenky G, Bliwise DL, Buxton OM, Buysse D, et al. Recommended Amount of Sleep for a Healthy Adult: A Joint Consensus Statement of the American Academy of Sleep Medicine and Sleep Research Society. Journal of Clinical Sleep Medicine. 2015;11(06):591–2. doi: doi:10.5664/jcsm.4758.

18. Nelson KL, Davis JE, Corbett CF. Sleep quality: An evolutionary concept analysis. Nurs Forum. 2022;57(1):144–51. Epub 20211005. doi: 10.1111/nuf.12659. PubMed PMID: 34610163.

19. Ohayon M, Wickwire EM, Hirshkowitz M, Albert SM, Avidan A, Daly FJ, et al. National Sleep Foundation’s sleep quality recommendations: first report. Sleep Health. 2017;3(1):6–19. Epub 20161223. doi: 10.1016/j.sleh.2016.11.006. PubMed PMID: 28346153.

20. Hinz A, Glaesmer H, Brähler E, Löffler M, Engel C, Enzenbach C, et al. Sleep quality in the general population: psychometric properties of the Pittsburgh Sleep Quality Index, derived from a German community sample of 9284 people. Sleep Medicine. 2017;30:57–63. doi: 10.1016/j.sleep.2016.03.008.

21. Kocevska D, Lysen TS, Dotinga A, Koopman-Verhoeff ME, Luijk MPCM, Antypa N, et al. Sleep characteristics across the lifespan in 1.1 million people from the Netherlands, United Kingdom and United States: a systematic review and meta-analysis. Nature Human Behaviour. 2021;5(1):113–22. doi: 10.1038/s41562-020-00965-x.

22. Kum-Nji P, Taylor S, Tanwi B. Doctor-diagnosed sleep disorders in the United States: Prevalence and impact of tobacco smoke exposure and vitamin D deficiency. A population-based study. Frontiers in Sleep. 2023;2. doi: 10.3389/frsle.2023.1113946.

23. Ahn E, Baek Y, Park J-E, Lee S, Jin H-J. Elevated prevalence and treatment of sleep disorders from 2011 to 2020: a nationwide population-based retrospective cohort study in Korea. BMJ Open. 2024;14(2):e075809. doi: 10.1136/bmjopen-2023-075809.

24. Schutte-Rodin S, Broch L, Buysse D, Dorsey C, Sateia M. Clinical guideline for the evaluation and management of chronic insomnia in adults. J Clin Sleep Med. 2008;4(5):487–504. PubMed PMID: 18853708; PubMed Central PMCID: PMCPMC2576317.

25. Limongi F, Siviero P, Trevisan C, Noale M, Catalani F, Ceolin C, et al. Changes in sleep quality and sleep disturbances in the general population from before to during the COVID-19 lockdown: A systematic review and meta-analysis. Frontiers in Psychiatry. 2023;14. doi: 10.3389/fpsyt.2023.1166815.

26. Zeng L-N, Zong Q-Q, Yang Y, Zhang L, Xiang Y-F, Ng CH, et al. Gender Difference in the Prevalence of Insomnia: A Meta-Analysis of Observational Studies. Frontiers in Psychiatry. 2020;11. doi: 10.3389/fpsyt.2020.577429.

27. Alimoradi Z, Gozal D, Tsang HWH, Lin C-Y, Broström A, Ohayon MM, et al. Gender-specific estimates of sleep problems during the COVID-19 pandemic: Systematic review and meta-analysis. Journal of Sleep Research. 2022;31(1):e13432. doi: 10.1111/jsr.13432.

28. Canever JB, Zurman G, Vogel F, Sutil DV, Diz JBM, Danielewicz AL, et al. Worldwide prevalence of sleep problems in community-dwelling older adults: A systematic review and meta-analysis. Sleep Med. 2024;119:118–34. Epub 20240405. doi: 10.1016/j.sleep.2024.03.040. PubMed PMID: 38669835.

29. Patel NP, Grandner MA, Xie D, Branas CC, Gooneratne N. "Sleep disparity" in the population: poor sleep quality is strongly associated with poverty and ethnicity. BMC Public Health. 2010;10(1):475. doi: 10.1186/1471-2458-10-475.

30. Johnson DA, Billings ME, Hale L. Environmental Determinants of Insufficient Sleep and Sleep Disorders: Implications for Population Health. Curr Epidemiol Rep. 2018;5(2):61–9. Epub 20180505. doi: 10.1007/s40471-018-0139-y. PubMed PMID: 29984131; PubMed Central PMCID: PMCPMC6033330.

31. Stenholm S, Head J, Kivimäki M, Magnusson Hanson LL, Pentti J, Rod NH, et al. Sleep Duration and Sleep Disturbances as Predictors of Healthy and Chronic Disease–Free Life Expectancy Between Ages 50 and 75: A Pooled Analysis of Three Cohorts. The Journals of Gerontology: Series A. 2018;74(2):204–10. doi: 10.1093/gerona/gly016.

32. Nistor P, Chang-Kit B, Nicholson K, Anderson KK, Stranges S. The relationship between sleep health and multimorbidity in community dwelling populations: Systematic review and global perspectives. Sleep Medicine. 2023;109:270–84. doi: 10.1016/j.sleep.2023.07.002.

33. Ohayon MM, Smirne S. Prevalence and consequences of insomnia disorders in the general population of Italy. Sleep Medicine. 2002;3(2):115–20. doi: 10.1016/S1389-9457(01)00158-7.

34. Léger D, Poursain B, Neubauer D, Uchiyama M. An international survey of sleeping problems in the general population. Current Medical Research and Opinion. 2008;24(1):307–17. doi: 10.1185/030079907X253771.

35. Varghese NE, Lugo A, Ghislandi S, Colombo P, Pacifici R, Gallus S. Sleep dissatisfaction and insufficient sleep duration in the Italian population. Scientific Reports. 2020;10(1):17943. doi: 10.1038/s41598-020-72612-4.

36. Sancho-Domingo C, Carballo JL, Coloma-Carmona A, Buysse DJ. Brief version of the Pittsburgh Sleep Quality Index (B-PSQI) and measurement invariance across gender and age in a population-based sample. Psychol Assess. 2021;33(2):111–21. Epub 20201029. doi: 10.1037/pas0000959. PubMed PMID: 33119375.

37. Buysse DJ, Reynolds CF, 3rd, Monk TH, Berman SR, Kupfer DJ. The Pittsburgh Sleep Quality Index: a new instrument for psychiatric practice and research. Psychiatry Res. 1989;28(2):193–213. doi: 10.1016/0165-1781(89)90047-4. PubMed PMID: 2748771.

38. Fabbri M, Beracci A, Martoni M, Meneo D, Tonetti L, Natale V. Measuring Subjective Sleep Quality: A Review. Int J Environ Res Public Health. 2021;18(3). Epub 20210126. doi: 10.3390/ijerph18031082. PubMed PMID: 33530453; PubMed Central PMCID: PMCPMC7908437.

39. Mollayeva T, Thurairajah P, Burton K, Mollayeva S, Shapiro CM, Colantonio A. The Pittsburgh sleep quality index as a screening tool for sleep dysfunction in clinical and non-clinical samples: A systematic review and meta-analysis. Sleep Med Rev. 2016;25:52–73. Epub 20150217. doi: 10.1016/j.smrv.2015.01.009. PubMed PMID: 26163057.

40. Curcio G, Tempesta D, Scarlata S, Marzano C, Moroni F, Rossini PM, et al. Validity of the Italian version of the Pittsburgh Sleep Quality Index (PSQI). Neurol Sci. 2013;34(4):511–9. Epub 20120413. doi: 10.1007/s10072-012-1085-y. PubMed PMID: 22526760.

41. Limesurvey GmbH. LimeSurvey: An Open Source survey tool Hamburg, Germany: LimeSurvey GmbH. Available from: http://www.limesurvey.org.

42. R Core Team. R: A Language and Environment for Statistical Computing. In: Computing RFfS, editor. Vienna, Austria: https://www.R-project.org/; 2023.

43. Posit team. RStudio: Integrated Development Environment for R. In: Posit Software P, editor. Boston, MA. : http://www.posit.co/; 2023.

44. Wickham, et al. Welcome to the Tidyverse. Journal of Open Source Software. 2019;4(43):1686,. doi: 10.21105/joss.01686.

45. Lumley T. Analysis of Complex Survey Samples. Journal of Statistical Software. 2004;9(8):1–19. doi: 10.18637/jss.v009.i08.

46. Bates D, Mächler M, Bolker B, Walker S. Fitting linear mixed-effects models using lme4 Journal of Statistical Software. 2015;67(1).

47. Filip I, Tidman M, Saheba N, Bennett H, Wick B, Rouse N, et al. Public health burden of sleep disorders: underreported problem. Journal of Public Health. 2017;25(3):243–8. doi: 10.1007/s10389-016-0781-0.

48. Perger E, Silvestri R, Bonanni E, Di Perri MC, Fernandes M, Provini F, et al. Gender medicine and sleep disorders: from basic science to clinical research. Frontiers in Neurology. 2024;15. doi: 10.3389/fneur.2024.1392489.

49. Edwards BA, O’Driscoll DM, Ali A, Jordan AS, Trinder J, Malhotra A. Aging and sleep: physiology and pathophysiology. Semin Respir Crit Care Med. 2010;31(5):618–33. Epub 20101012. doi: 10.1055/s-0030-1265902. PubMed PMID: 20941662; PubMed Central PMCID: PMCPMC3500384.

50. Berkley AS, Carter PA, Yoder LH, Acton G, Holahan CK. The effects of insomnia on older adults’ quality of life and daily functioning: A mixed-methods study. Geriatric Nursing. 2020;41(6):832–8. doi: 10.1016/j.gerinurse.2020.05.008.

51. Chu Y, Oh Y, Gwon M, Hwang S, Jeong H, Kim H-W, et al. Dose-response analysis of smartphone usage and self-reported sleep quality: a systematic review and meta-analysis of observational studies. Journal of Clinical Sleep Medicine. 2023;19(3):621–30. doi: doi:10.5664/jcsm.10392.

52. Johnson DA, Jackson CL, Williams NJ, Alcántara C. Are sleep patterns influenced by race/ethnicity - a marker of relative advantage or disadvantage? Evidence to date. Nat Sci Sleep. 2019;11:79–95. Epub 20190723. doi: 10.2147/nss.S169312. PubMed PMID: 31440109; PubMed Central PMCID: PMCPMC6664254.

53. Li J, Cao D, Huang Y, Chen Z, Wang R, Dong Q, et al. Sleep duration and health outcomes: an umbrella review. Sleep Breath. 2022;26(3):1479–501. Epub 20210826. doi: 10.1007/s11325-021-02458-1. PubMed PMID: 34435311.

54. Kemple M, O’Toole S, O’Toole C. Sleep quality in patients with chronic illness. J Clin Nurs. 2016;25(21-22):3363–72. Epub 20160815. doi: 10.1111/jocn.13462. PubMed PMID: 27378192.

55. Holder S, Narula NS. Common Sleep Disorders in Adults: Diagnosis and Management. Am Fam Physician. 2022;105(4):397–405. PubMed PMID: 35426627.

56. Wiedermann CJ, Sangermano K, Marino P, Ausserhofer D, Engl A, Piccoliori G. Distinct Regional Pattern of Sedative Psychotropic Drug Use in South Tyrol: A Comparison with National Trends in Italy. Preprints: Preprints; 2024.

57. Baranwal N, Yu PK, Siegel NS. Sleep physiology, pathophysiology, and sleep hygiene. Progress in Cardiovascular Diseases. 2023;77:59–69. doi: 10.1016/j.pcad.2023.02.005.

58. Dunietz GL, Jansen EC, Chervin RD. What Should a Public Health Approach to Sleep Look Like? AMA J Ethics. 2024;26(10):E795–803. Epub 20241001. doi: 10.1001/amajethics.2024.795. PubMed PMID: 39361393.

59. SIivertsen B, Nordhus IH, Bjorvatn B, Pallesen S. Sleep problems in general practice: a national survey of assessment and treatment routines of general practitioners in Norway. Journal of Sleep Research. 2010;19(1-Part-I):36-41. doi: 10.1111/j.1365-2869.2009.00769.x.

